# The global prevalence and ethnic heterogeneity of iron-refractory iron deficiency anaemia

**DOI:** 10.1101/2022.05.31.22275755

**Authors:** Shanghua Fan, Ting Zhao, Liu Sun

## Abstract

**Background:** Iron-refractory iron deficiency anaemia (IRIDA) is an autosomal recessive iron deficiency anaemia caused by mutations in the *TMPRSS6* gene. Iron deficiency anaemia is common, whereas IRIDA is rare. The prevalence of IRIDA is unclear. This study aimed to estimate the carrier frequency and genetic prevalence of IRIDA using Genome Aggregation Database (gnomAD) data.

**Methods:** The pathogenicity of *TMPRSS6* variants was interpreted according to the American College of Medical Genetics and Genomics (ACMG) and the Association for Molecular Pathology (AMP) standards and guidelines. The minor allele frequency (MAF) of *TMPRSS6* gene disease-causing variants in 141456 unique individuals was examined to estimate the global prevalence of IRIDA in seven ethnicities: African/African American (afr), American Admixed/Latino (amr), Ashkenazi Jewish (asj), East Asian (eas), Finnish (fin), Non-Finnish European (nfe) and South Asian (sas). The global and population-specific carrier frequencies and genetic prevalence of IRIDA were calculated using the Hardy-Weinberg equation.

**Results:** In total, 86 pathogenic/likely pathogenic variants (PV/LPV) were identified according to ACMG/AMP guideline. The global carrier frequency and genetic prevalence of IRIDA were 2.02 per thousand and 1.02 per million, respectively.

**Conclusions:** The prevalence of IRIDA is greater than previous estimates.

## Background

Iron-refractory iron deficiency anaemia (IRIDA) (OMIM: 206200, ORPHA: 209981) is a refractory iron deficiency anaemia that is unresponsive to oral iron treatment[1-4], leading to microcytic hypochromic anaemia, low transferrin saturation and serum iron[5]. Symptoms of IRIDA are usually mild, including tiredness (fatigue), weakness, pale skin, dizziness, and exercise-induced dyspnoea. These symptoms are most pronounced during childhood. Most IRIDA patients have normal growth and development. [5] IRIDA is usually suspected in childhood based on the results of routine blood tests. Diagnosis is according to laboratory tests showing hypochromic microcytic anaemia with low serum iron and transferrin saturation levels, and then the diagnosis is confirmed by genetic testing[6]. Treatment usually involves intravenous (IV) iron therapy[7].

IRIDA is a rare autosomal recessive disorder caused by mutations in the *TMPRSS6* (transmembrane serine protease 6) gene [8]. The gene encodes a serine protease that plays an essential role in downregulating hepcidin, the key regulator of iron homeostasis[9-11], through the cleavage of the cell surface haemojuvelin BMP coreceptor (*HJV*), an activator of hepcidin expression [12-15]. The *TMPRSS6* gene, located on chromosome 22, spans 18 exons and comprises 51125 base pairs, encoding a protein of 802 amino acids. Since 2008, more than 100 *TMPRSS6* variants have been identified in patients with IRIDA[16-18]. (The full paper list and full variant list are available in the Science Data Bank (ScienceDB) repository)

As high-throughput sequencing technology has evolved, re-evaluation guidelines for interpreting and classifying the pathogenicity of identified variants have been implemented. In addition, large-scale population databases have become widely available and can be used for assessment of genetic variants in rare diseases. In fact, for several diseases, there is evidence that these databases have improved the interpretation and classification of variants in patients with monogenic disease and allowed better prediction of which variants are likely to cause disease. The most widely used large-scale reference dataset is from Genome Aggregation Database (gnomAD), which consists of exome sequencing and genome sequencing data from unrelated individuals of diverse ancestries. [19]

The World Health Organization (WHO) defines anaemia as a condition in which the number of red cells is insufficient to meet the body’s physiological needs. Iron deficiency anaemia (IDA) is the most common cause of anaemia worldwide. Although iron deficiency anaemia is relatively common[3], IRIDA is rare. Although its prevalence is unknown, the Orphanet database (https://www.orpha.net/consor/cgi-bin/index.php) estimates is to be less than 1 per 1 million. We attempted to obtain a more reliable estimate of the prevalence and carrier frequency of IRIDA from the Genome Aggregation Database (gnomAD) dataset using a previous protocol[20]. Additionally, we aimed to generate a curated artificial intelligence training dataset of *TMPRSS6* variants for pathogenicity classification and interpretation.

## Methods

### Identification of previously reported *TMPRSS6* disease-causing variants

To evaluate the genetic spectrum of IRIDA, a comprehensive search was performed to identify all previously reported disease-causing *TMPRSS6* gene variants. Searches were conducted in the PubMed or Scopus database using the following combinations of search terms: iron deficiency anaemia, iron deficiency, iron-refractory iron deficiency anaemia, iron-refractory iron deficiency anaemia, Matriptase-2 (MT2), *tmprss6*, transmembrane serine protease 6, IRIDA, mutations, variants, variations, and mutants.

Two independent authors screened publications according to inclusion and exclusion criteria: original case reports reporting disease-causing variants of the *TMPRSS6* gene were included, and variants in abstract, full-text, tables, figures, or supplementary figures and tables were extracted. Non-English-language articles, reviews, comments, editorials, letters, etc., and *in vitro* and animal model studies were excluded. Common *TMPRSS6* polymorphisms associated with iron deficiency/iron deficiency anaemia in genome-wide association studies were also excluded.

All publications were saved in Medline format and stored in the MySQL and MongoDB databases using NCBI Entrez Programming Utilities [17] (E-utilities) with the Python package biopython [18] and MySQL/MongoDB database implementation. Reported *TMPRSS6* variants were also identified from Leiden Open Variation Database (LOVD) (https://www.lovd.nl/), NCBI[21] ClinVar (https://www.ncbi.nlm.nih.gov/clinvar/), dbSNP (https://www.ncbi.nlm.nih.gov/snp/), VarSome[22] (https://varsome.com/), Ensembl [23] (https://www.ensembl.org/) UniProtKB[24] (https://www.uniprot.org/), Genomenon Mastermind[25] (https://mastermind.genomenon.com/), LitVar [26] (https://www.ncbi.nlm.nih.gov/research/litvar2/), Online Mendelian Inheritance in Man (OMIM) (https://www.omim.org/) and HGMD (http://www.hgmd.cf.ac.uk/ac/index.php).

Prediction and functional annotation for all *TMPRSS6* potential nonsynonymous single-nucleotide variants (nsSNVs) were compiled using dbNSFP 4. [27]

### Identification and prediction of novel *TMPRSS6* disease-causing variants

The gnomAD database (https://gnomad.broadinstitute.org/) was searched for novel disease-causing *TMPRSS6* variants that had not yet been reported, and protein-truncating variants (PTVs) were examined (frameshifts, stop codons, initiator codons, splice donors and splice acceptors).

### Interpretation of the pathogenicity of *TMPRSS6* variants

The pathogenicity of variants was interpreted following the protocol by Zhang *et al*.[28] The pathogenicity of all missense, synonymous, and protein-truncating *TMPRSS6* variants was classified according to Standards and Guidelines of the American College of Medical Genetics and Genomics (ACMG) and the Association for Molecular Pathology (AMP) [29] with the ClinGen Variant Curation Interface[30]. For missense and synonymous variants, we performed additional literature retrieval to curate *in vitro* or *in vivo* functional studies supportive of a damaging effect on *TMPRSS6* missense/synonymous variants.

Variants classified as pathogenic and likely pathogenic were included. Variants classified as benign, likely benign, or uncertain significance were excluded. Pathogenic/likely pathogenic variants were included in the carrier frequency and genetic prevalence calculation.

### Annotation of variants with minor allele frequencies (MAFs)

Each identified variant was normalized with HGVS nomenclature, SPDI [31], VCF, ClinVar, ClinGen Allele Registry[32], and dbSNP Reference SNP (rs or RefSNP) using Mutalyzer[33] (https://mutalyzer.nl/), NCBI Entrez Programming Utilities (E-utilities), VariantValidator[34] (https://variantvalidator.org/) and Ensembl REST API (https://rest.ensembl.org/).

The canonical *TMPRSS6* transcript (NM_001374504.1, 3,265 bp) and protein (NP_001361433.1, 802 aa) selected by Matched Annotation from NCBI and EMBL-EBI (MANE) [35] differ from the 811 aa isoform in the literature. The difference between the two isoforms is that translation is initiated at an alternate start codon <u>M</u>LLLFHSKR<u>M</u>. [36].

gnomAD v2.1.1 VCF files in Parquet format were downloaded from Microsoft Azure Open Datasets with the Microsoft AzCopy tool. The minor allele frequency of the gnomAD population for the following ethnic groups were retrieved with Apache Drill, Apache Spark, and Apache Zeppelin: African/African American (AFR), American Admixed/Latino (AMR), East Asian (EAS), Non-Finish European (NFE) and South Asian (SAS).

### Carrier frequency and genetic prevalence calculation

The carrier frequency and genetic prevalence of IRIDA were calculated based on the Hardy–Weinberg equation[37]. For a monogenic autosomal recessive disorder, the genetic prevalence is [1 - ∏_*i*_(1 - *q*_*i*_)]^2^ based on the theory of probability, where q_i_ stands for each pathogenic/likely pathogenic variant minor allele frequency. The genetic prevalence was approximately equal to (∑ *q*_*i*_)^2^ and the carrier frequency was 2(1 - ∑ *q*_*i*_) ∑ *q*_*i*_ ≈ 2 ∑ *q*_*i*_ . The disease prevalence was estimated by using the observed allele frequency of a pathogenic/likely pathogenic variant in the gnomAD database as the direct estimator for q_i_. The disease prevalence can be estimated by 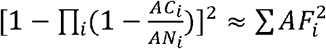, where AC is the allele count, AN is the corresponding allele number and AF is the corresponding allele frequency.

The Python statistics package statsmodels and scientific computing package NumPy and pandas were employed for calculating the 95% confidence interval (95% CI) for the binomial proportion of carrier frequency and the genetic prevalence with the Wilson Score interval. Graphics were plotted using the R graphic package ggplot2.

## Results

### Identification of *TMPRSS6* missense, synonymous and protein-truncating variants

Comprehensive retrieval of IRIDA disease-causing variants resulted in the identification of 813 articles, of which 39 were considered eligible according to the exclusion and inclusion criteria. All disease-causing variant ETL pipelines and tools are shown in Fig. 1. From these articles, 86 disease-causing variants in the *TMPRSS6* gene, including 40 missense variants and 46 protein-truncating variants (PTVs), are classified as pathogenic/likely pathogenic according to the Standards and guidelines of the American College of Medical Genetics and Genomics and the Association for Molecular Pathology (missense variants are shown in Table 1; all variants are listed in the Science Data Bank (ScienceDB) online data repository). The most common mutation consequence is missense, accounting for 46.5% (40/86) of all pathogenic/likely pathogenic variants and more than half of the total allele frequency.

**Table 1.**
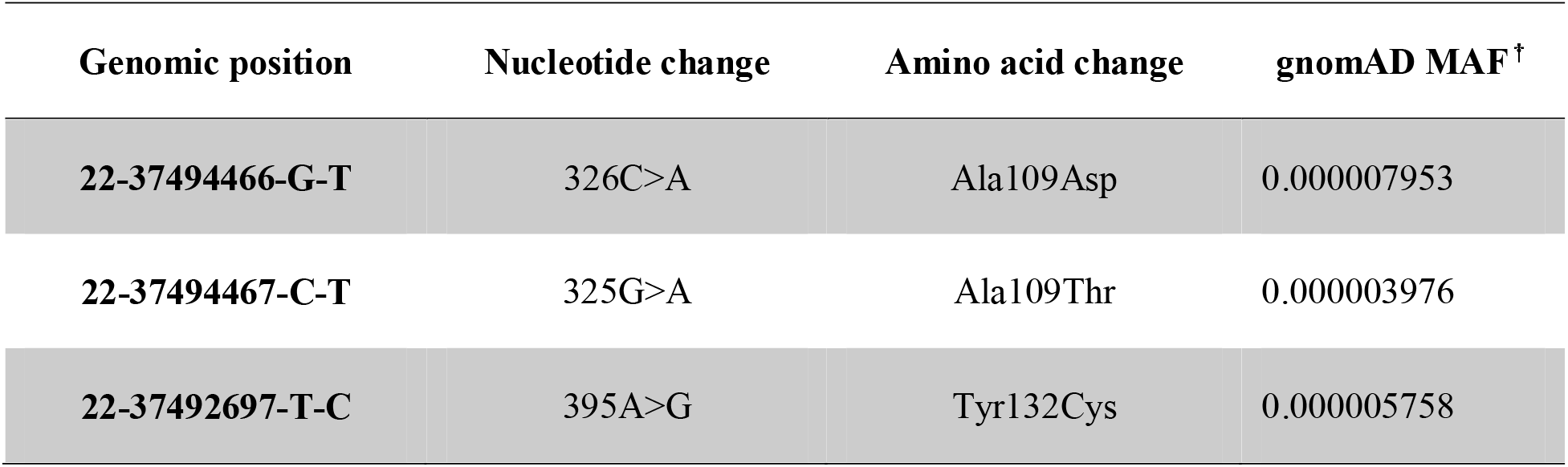
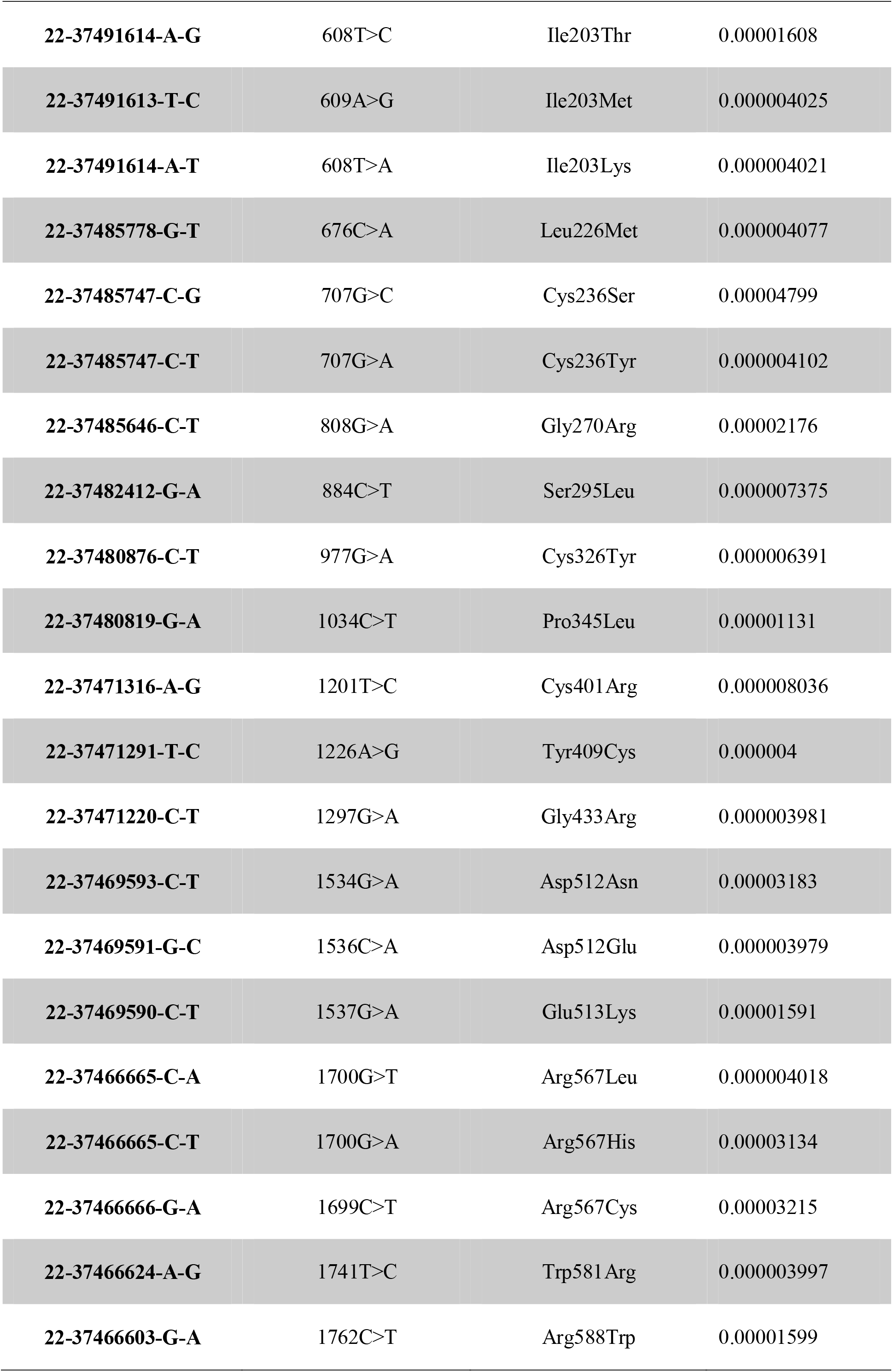
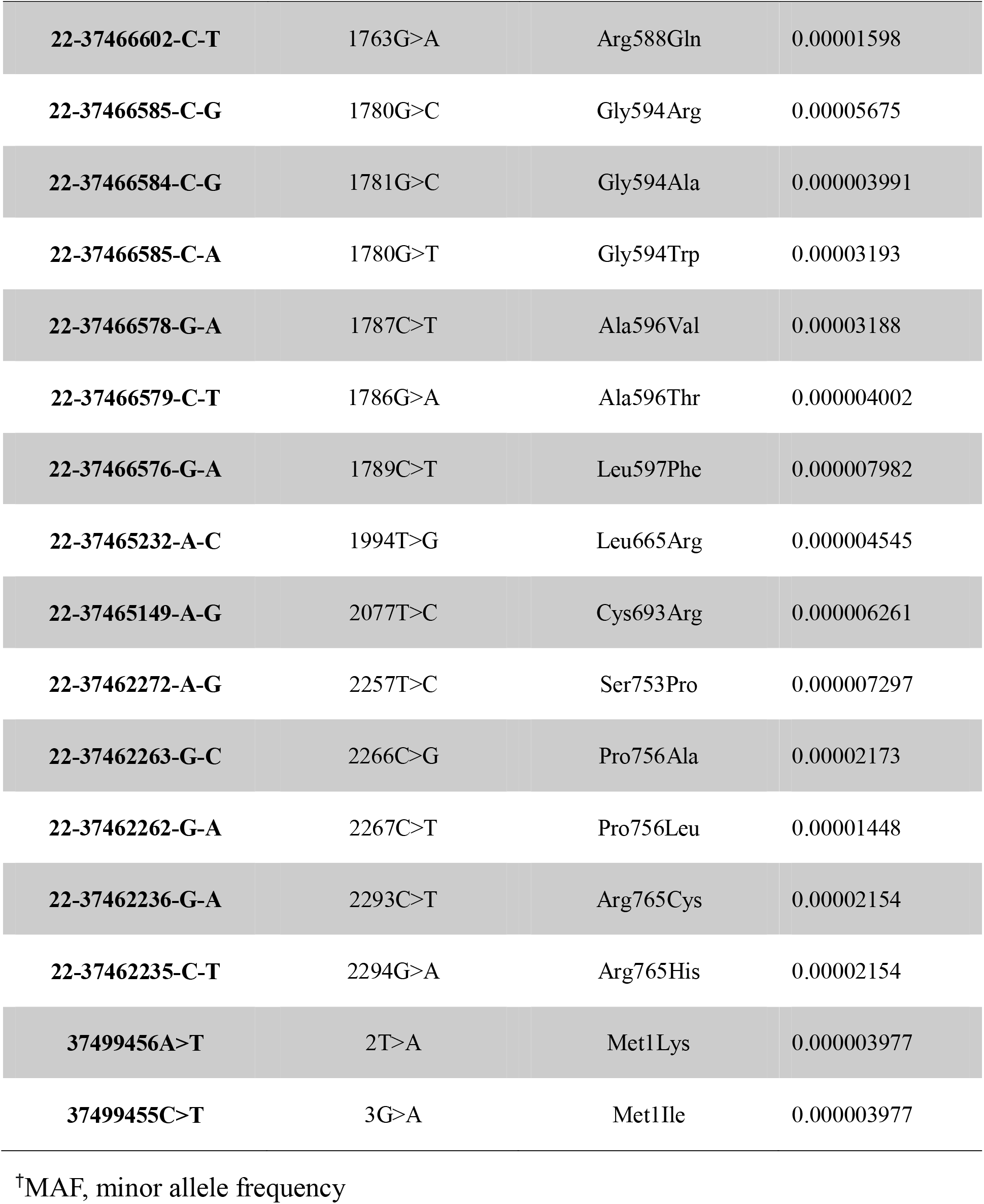
*TMPRSS6* pathogenic/likely pathogenic missense variants present in gnomAD with global minor allele frequencies

**Figure 1.**
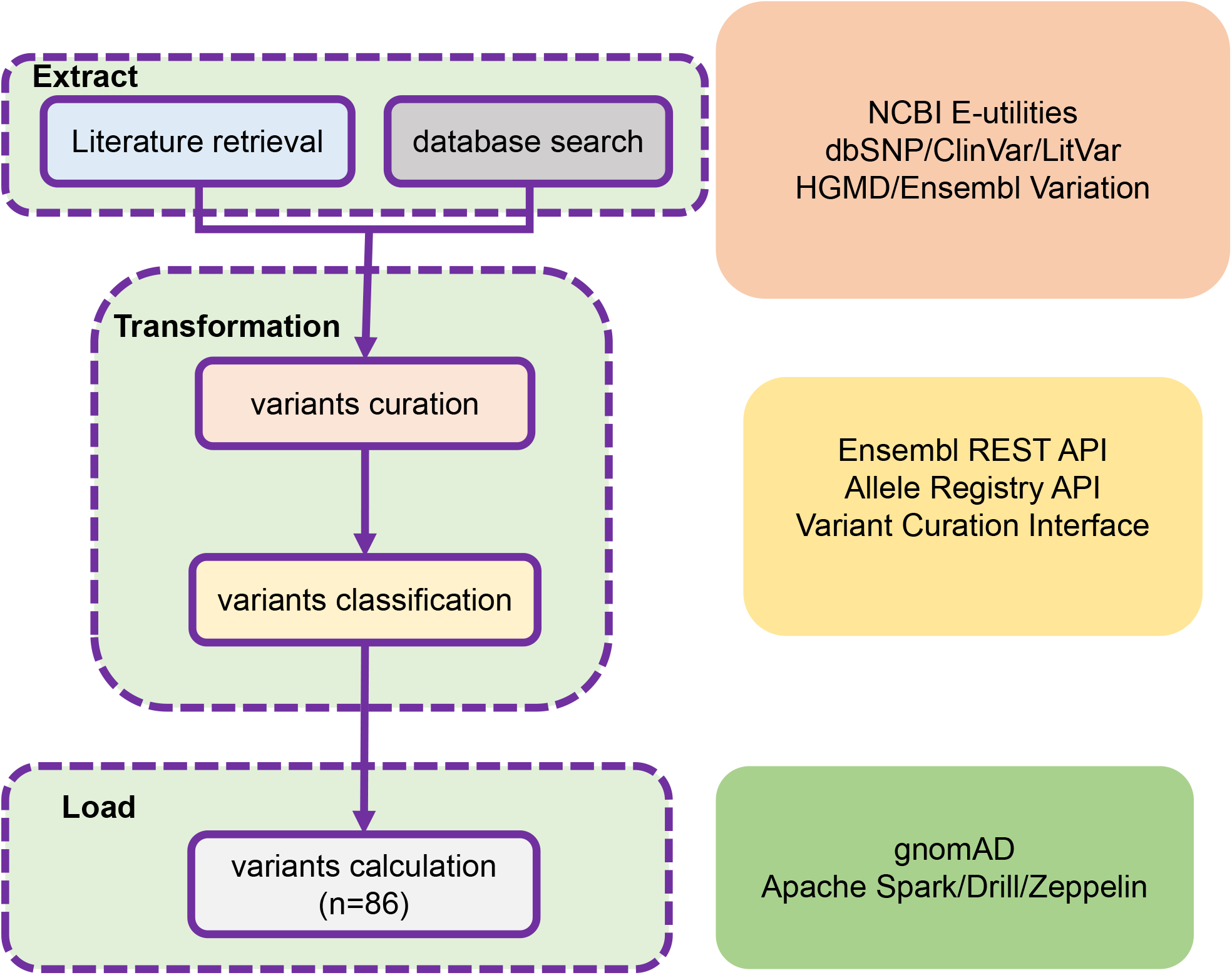
ETL pipeline of curation and classification of TMPRSS6 pathogenic/likely pathogenic variants. **Extract**: variants from literature retrieval and database search. **Transformation**: raw variant normalized HGVS nomenclature and curated. **Load**: based on the Hardy-Weinberg equation and binomial distribution model calculation

One synonymous variant (c.1086G>A/c.1086G>T, p.Thr362=) is classified as likely pathogenic [38] (PP3, PP4, PM2, and PS3 criteria). However, this variant was excluded from the gnomAD dataset because of low confidence pLoF.

Forty-six protein-truncating variants were divided into three categories: frameshift (n=20), nonsense (n=17), and splice acceptor/donor (n=9). PTVs accounted for more than half of the number of pathogenic/likely pathogenic variants after filtering (low confidence pLoF, LCR, Not LoF) but less than half of the total allele frequency.

### Estimation of population prevalence and the carrier frequency of IRIDA

Pooling of the allele frequencies for all pathogenic/likely pathogenic variants provided a global minor allele frequency of 0.001, which is equivalent to a genetic prevalence of 1.0228 per 1 million (95% CI: [0.82, 1.28]) and a carrier frequency of 2.02 per 1 thousand (95% CI: [1.8, 2.28]). The African population had the highest prevalence, at 3.55 per million (95% CI: [1.76, 7.18]), with carrier frequency at 3.77 per thousand. Both non-Finish European and East Asian populations had a prevalence of greater than 1 per million, whereas the prevalence of American/Admixed Latino and South Asian populations was smaller than 1 per million. Ashkenazi Jewish and Finnish populations were not analysed because the sample size was too small to be calculated (Table 2 and Fig. 2).

**Table 2.** Estimated prevalence and carrier frequency among ethnicities and ancestries.

| Population | Genetic prevalence |  | Carrier frequency |  |
| --- | --- | --- | --- | --- |
|  | (per million) |  | (per thousand) |  |
|  |  | 95% CI |  | 95% CI† |
| Non-Finnish European | 1.54 | 0.87, 2.5 | 2.48 | 2.11, 2.92 |
| American Admixed/Latino | 0.14 | 0.05, 0.41 | 0.75 | 0.44, 1.29 |
| African/African American | 3.55 | 1.76, 7.18 | 3.77 | 2.65, 5.36 |
| **South Asian** | 0.75 | 0.35, 1.6 | 1.73 | 1.19, 2.53 |
| **East Asian** | 1.51 | 0.67, 3.42 | 2.46 | 0.69, 2.18 |
| **All** | 1.02 | 0.82, 1.28 | 2.02 | 1.8, 2.28 |

†CI: confidence interval

**Fig. 2.**
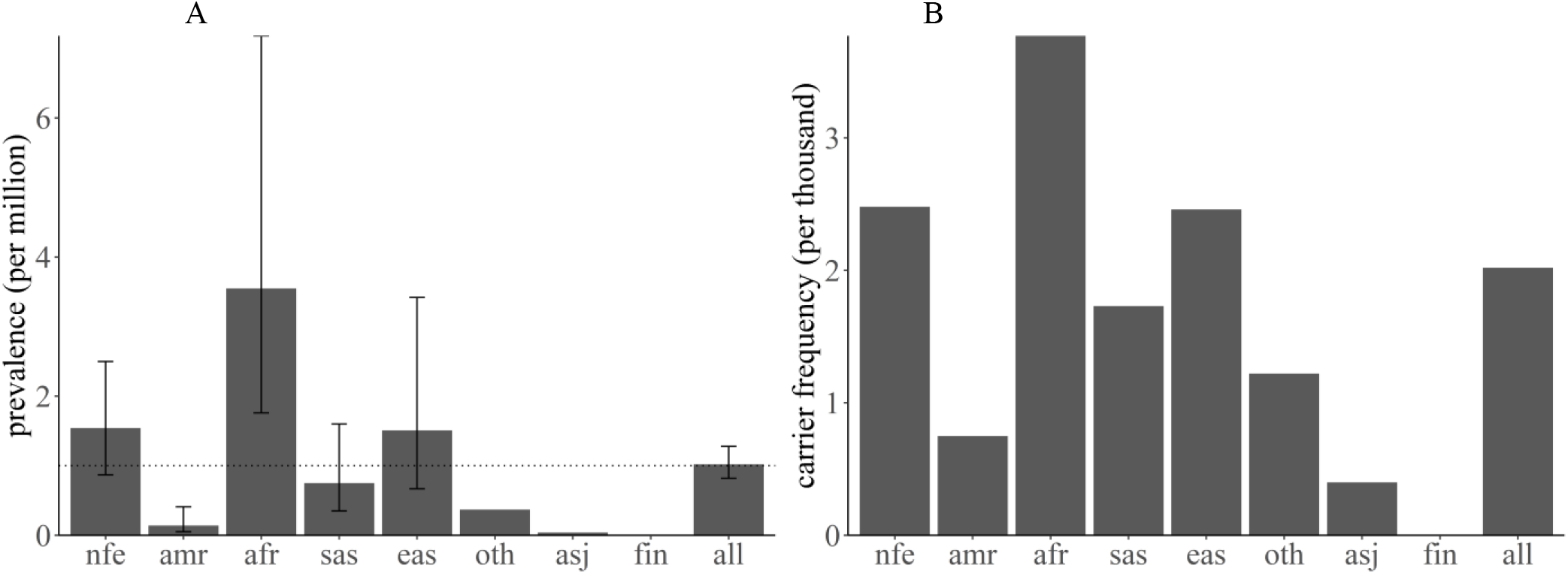
genetic prevalence and carrier frequency of IRIDA. **A**. IRIDA genetic prevalence estimated from gnomAD allele frequencies. B. IRIDA carrier frequency among diverse population. African/African American (afr), American Admixed/Latino (amr), Ashkenazi Jewish (asj), East Asian (eas), Finnish (fin), Non-Finnish European (nfe) and South Asian (sas), Other(oth)

## Discussion

IRIDA is a rare autosomal recessive disorder caused by homozygous or compound heterozygous mutations in the transmembrane serine protease 6 (*TMPRSS6*) gene. IRIDA patients have microcytic hypochromic anaemia, low serum iron, and transferrin saturation. The prevalence is unknown. In this study, we sought to estimate the prevalence of IRIDA using 100k-scale population genome data and to deepen our understanding of *TMPRSS6* genetic variation. Using gnomAD data, we found that IRIDA affects 1.0228 per 1 million in the global population, which is higher than previous Orphanet database estimates of less than 1 per 1 million.

The main strength of this study was the use of 86 *TMPRSS6* variants from a population of more than 100 k for prevalence estimates. In addition, we produced a curated variant dataset for *TMPRSS6* that will be useful for further artificial intelligence-based classification of germline variant pathogenicity.

IRIDA can be difficult to distinguish from acquired iron deficiency unresponsive to oral iron therapy. Overall, IRIDA is a genotypically and phenotypically heterogeneous disease. Allelic *TMPRSS6* mutations are found in most patients with IRIDA. An algorithm has been developed based on blood and plasma values to distinguish patients with IRIDA resulting from biallelic *TMPRSS6* mutations [40-42].

Combined heterozygous mutations in *TMPRSS6* and activin A receptor type 1 (ACVR1) have been reported to cause IRIDA[43]. High hepcidin expression in the proband was reported with ACVR1 R258S and *TMPRSS6* I212T. This digenic model suggests that combined heterozygous mutations in *TMPRSS6* and other BMP/SMAD signalling pathway genes are likely to cause IRIDA. This may explain index cases in which there is only one pathogenic mutation and one or more single-nucleotide polymorphism combination.

*TMPRSS6* polymorphisms are more frequent than germline *TMPRSS6* mutations, and common *TMPRSS6* variants are associated with erythrocyte traits, including haemoglobin concentration (Hb), haematocrit (Hct), mean corpuscular volume (MCV), mean corpuscular haemoglobin (MCH), mean corpuscular haemoglobin concentration (MCHC) and red blood cell (RBC) count[44, 45]. Additionally, *TMPRSS6* polymorphisms in iron deficiency anaemia are partially responsive to oral iron treatment[46, 47]. A 3-tier classification system for variants of the *TMPRSS6* gene has been proposed to better understand and define the IRIDA spectrum. [48] Synonymous mutations are also called neutral or silent mutations. Synonymous mutations in protein-coding genes do not alter protein sequences and are thus generally presumed to be neutral. Shen *et al*. constructed 8,341 yeast mutants and measured their fitness relative to wildtype. They found that three-quarters of synonymous mutations resulted in a significant reduction in fitness; the distribution of fitness effects was overall similar, albeit nonidentical[49].

IRIDA is not on the first national list of rare diseases issued by China, and the prevalence of IRIDA in China has not been estimated. [50] The National Rare Diseases Registry System (NRDRS) has not registered any IRIDA cases. [51] We demonstrated the power and limitations of the 100k population genome database [52, 53] to calculate the prevalence of rare diseases, but gnomAD and the Trans-Omics for Precision Medicine (TOPMed) population are of predominantly non-Asian ancestry. The Chinese population genome project will fill this gap. Currently, NyuWa[54], Westlake BioBank for Chinese (WBBC)[55], and PGG. Han[56], China Metabolic Analytics Project (ChinaMAP)[57] have been released. We will estimate the prevalence of IRIDA and other rare diseases through the aggregation of Chinese population genome data.

The number of *TMPRSS6* curated variants in the ClinVar database is far less than the number of variants we manually curated, and the state-of-the-art model EVE (evolutionary model of variant effect) uses the ClinVar dataset as the training set[58]. ClinGen Variant Curation Expert Panels (VCEPs) [59] define biocuration application of ACMG/AMP guidelines for sequence variant interpretation for specific genes or diseases. Additionally, the ClinGen Variant Curation Interface [30] is a variant classification platform for the application of evidence criteria from ACMG/AMP guidelines for curating variant classifications.

In conclusion, through a comprehensive analysis of genetic variation in *TMPRSS6*, we expanded our recognition of disease-causing mutations to 86 variants. IRIDA is a rare disease, and these data can help clinicians to diagnose IRIDA that is unlikely to represent a significant proportion of patients presenting with refractory iron deficiency or iron-deficiency anaemia.

## Data Availability

The datasets are available online in the Science Data Bank (ScienceDB) repository.

https://doi.org/10.11922/sciencedb.01515

## Abbreviations

TMPRSS6: transmembrane serine protease 6
IRIDA: iron refractory iron deficiency anaemia
MAF: minor allele frequency
ClinGen: Clinical Genome
NCBI: National Center for Biotechnology Information
ACMG: American College of Medical Genetics and Genomics
AMP: Association for Molecular Pathology
VCEP: Variant Curation Expert Panel
CI: confidence interval
gnomAD: Genome Aggregation Database
ID: iron deficiency
IDA: iron deficiency anaemia
PTVs: protein-truncating variants
AC: allele count
AF: allele frequency
AN: allele number

## Declarations

### Ethics approval and consent to participate

Not applicable.

### Consent for publication

Not applicable.

### Availability of data and materials

The datasets are available online in the Science Data Bank (ScienceDB) repository. https://doi.org/10.11922/sciencedb.01515

### Author Contributions

FSH and ZT retrieved the literature and wrote the manuscript text, and SL designed the project and revised the manuscript and data analysis. All authors read and approved the final manuscript.

## Acknowledgements

We acknowledge Genome Aggregation Database (gnomAD).

